# COVID-19-related mortality by age groups in Europe: A meta-analysis

**DOI:** 10.1101/2020.04.11.20061721

**Authors:** Jérémie F. Cohen, Daniël A. Korevaar, Soraya Matczak, Joséphine Brice, Martin Chalumeau, Julie Toubiana

**Affiliations:** Université de Paris, Centre of Research in Epidemiology and Statistics - CRESS, INSERM, F-75004 Paris, France; Department of General Pediatrics and Pediatric Infectious Diseases, AP-HP, Necker Hospital for Sick Children, Université de Paris, 75015, Paris, France; Department of Respiratory Medicine, Amsterdam University Medical Center, University of Amsterdam, PO Box 22700, 1100 DE, Amsterdam, The Netherlands

**Keywords:** Coronavirus, SARS-CoV-2, COVID-19, Mortality, Age, Meta-analysis

## Abstract

**Background and Objectives:** To date, more than 1,000,000 confirmed cases and 65,000 deaths due to coronavirus disease 2019 (COVID-19) have been reported globally. Early data have indicated that older patients are at higher risk of dying from COVID-19 than younger ones, but precise international estimates of the age-breakdown of COVID-19-related deaths are lacking.

**Materials and Methods:** We evaluated the distribution of COVID-19-related fatalities by age groups in Europe. On April 6, 2020, we systematically reviewed COVID-19-related mortality data from 32 European countries (European Union/European Economic Area and the United Kingdom). We collated official reports provided by local Public Health or Ministry of Health websites. We included countries if they provided data regarding more than 10 COVID-19-related deaths stratified by age according to pre-specified groups (i.e., < 40, 40-69, ≥ 70 years). We used random-effects meta-analysis to estimate the proportion of age groups among all COVID-19-related fatalities.

**Results:** Thirteen European countries were included in the review, for a total of 31,864 COVID-19-related deaths (range: 27-14,381 per country). In the main meta-analysis (including data from Germany, Hungary, Italy, Netherlands, Portugal, Spain, Switzerland; 21,522 COVID-19-related fatalities), the summary proportions of persons < 40, 40-69, and ≥ 70 years of age among all COVID-19-related deaths were 0.1% (0.0-0.2%; *I*^*2*^ 24%), 12.8% (10.3-15.6%; *I*^*2*^ 94%), and 84.8% (81.3-88.1%; *I*^*2*^ 96%), respectively.

**Conclusions:** People under 40 years of age represent a small fraction of the total number of COVID-19-related deaths in Europe. These results may help health authorities respond to public concerns and guide future physical distancing and mitigation strategies.

## Introduction

As of April 6, 2020, more than 1,000,000 confirmed cases and 65,000 deaths due to coronavirus disease 2019 (COVID-19) have been reported globally [1]. Data from China and Italy have indicated that older adults are at higher risk of dying from COVID-19 than are younger persons [2, 3]. However, to date, most studies have emphasized the case-fatality rate [3-5], i.e., the conditional probability of death among those classified as COVID-19 cases. This indicator is likely to be biased in the early phase of an outbreak, mostly because of preferential testing of those with more severe disease (e.g., hospitalized patients) and delay between the time of death and its official registration [6].

While there is a growing sense that SARS-CoV-2 can result in severe disease regardless of age [7], precise estimates of the age-breakdown of COVID-19-related deaths are lacking. Here we evaluated the distribution of COVID-19-related fatalities by age in Europe, the most impacted region in the world. Such an analysis is critical to direct any potential relaxation of physical distancing and mitigation measures that are highly effective in reducing the epidemic, but have severe economic and social consequences [8]. We hypothesized that people less than 40 years of age represent a small fraction of all COVID-19-related deaths, as suggested by early reports from the Chinese Centers for Disease Control and Prevention (CDC) [2].

## Materials and Methods

On April 6, 2020, one of the authors (J.F.C.) systematically reviewed and extracted COVID-19-related mortality data from all 32 European countries participating in the European CDC surveillance network (i.e., European Union/European Economic Area and the United Kingdom), by collating official reports provided by local Public Health or Ministry of Health websites (Appendix 1). A second author (D.A.K.) verified all data extraction, and disagreements were discussed. We included countries if they provided data regarding more than 10 COVID-19-related deaths stratified by age according to pre-specified groups (i.e., < 40, 40-69, ≥ 70 years). We applied no language restrictions.

We used random-effects meta-analysis to estimate the proportion of age groups (< 40, 40-69, ≥ 70 years) among all COVID-19-related fatalities. Heterogeneity was quantified with the *I*^*2*^ statistic. We conducted a sensitivity analysis that also included countries that reported data using slightly different age groups, with the following inclusion criteria: (1) the cutoff to define the ‘younger’ age group lied within 40-50; (2) the ‘intermediate’ age group covered at least 20 years (e.g., UK, < 40, 40-59, ≥ 60 years). Statistical analyses involved use of Stata 15/SE (Stata Corp., College Station, Texas).

## Results

We identified official reports on COVID-19-related mortality data for all European countries except Cyprus, for a total of 32,977 fatalities. Eighteen countries (accounting for 1,113 deaths) were excluded because of unusable data: eleven were excluded because of the lack of age-breakdown of the COVID-19 deaths, five because of fewer than 10 fatalities, and two because of age cutoffs not matching our pre-defined groups (Appendix 1). Thirteen European countries were included in the review, for a total of 31,864 COVID-19-related deaths (range: 27-14,381 per country; Table 1).

**Table 1.**
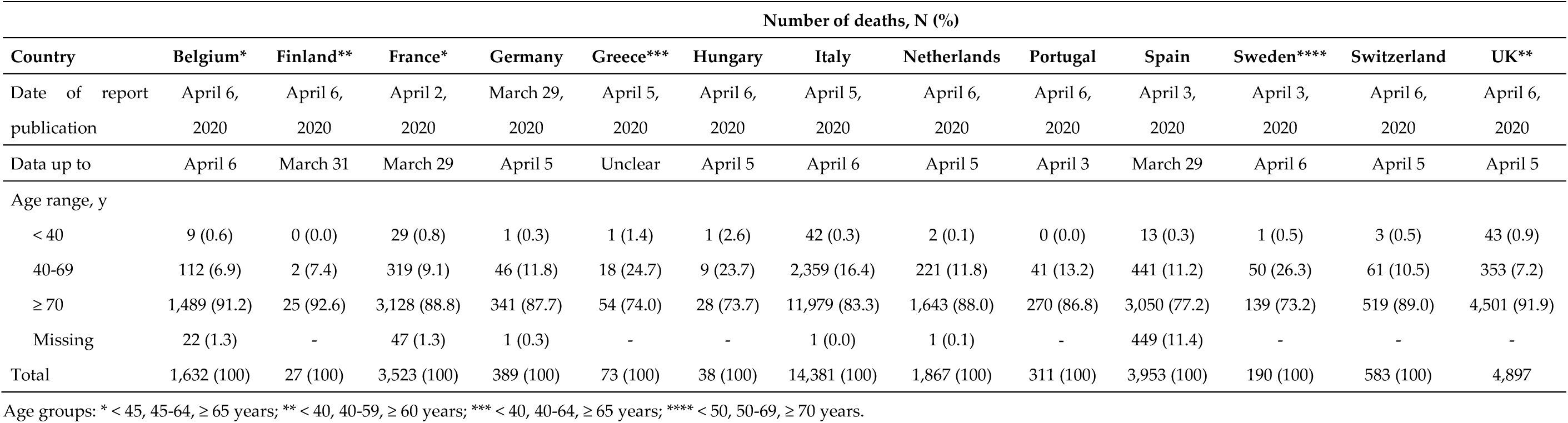
Death by COVID-19 by age groups in Europe (as of April 6, 2020)

In the main meta-analysis (7 countries; 21,522 COVID-19-related fatalities), the summary proportions of persons < 40, 40-69, and ≥ 70 years of age among all COVID-19-related deaths were 0.1% (0.0-0.2%; *I*^*2*^ 24%), 12.8% (10.3-15.6%; *I*^*2*^ 94%), and 84.8% (81.3-88.1%; *I*^*2*^ 96%), respectively (Figure 1). In a sensitivity analysis also including countries that reported slightly different age groups (13 countries; 31,864 COVID-19-related fatalities), the summary proportions of persons around < 40-50, around 40-69, and around ≥ 60-70 years of age among all COVID-19-related deaths were 0.2% (0.1-0.4%; *I*^*2*^ 75%), 12.5% (9.8-15.5%; *I*^*2*^ 98%), and 85.5% (82.1-88.6%; *I*^*2*^ 98%), respectively (Appendix 2).

**Figure 1.**
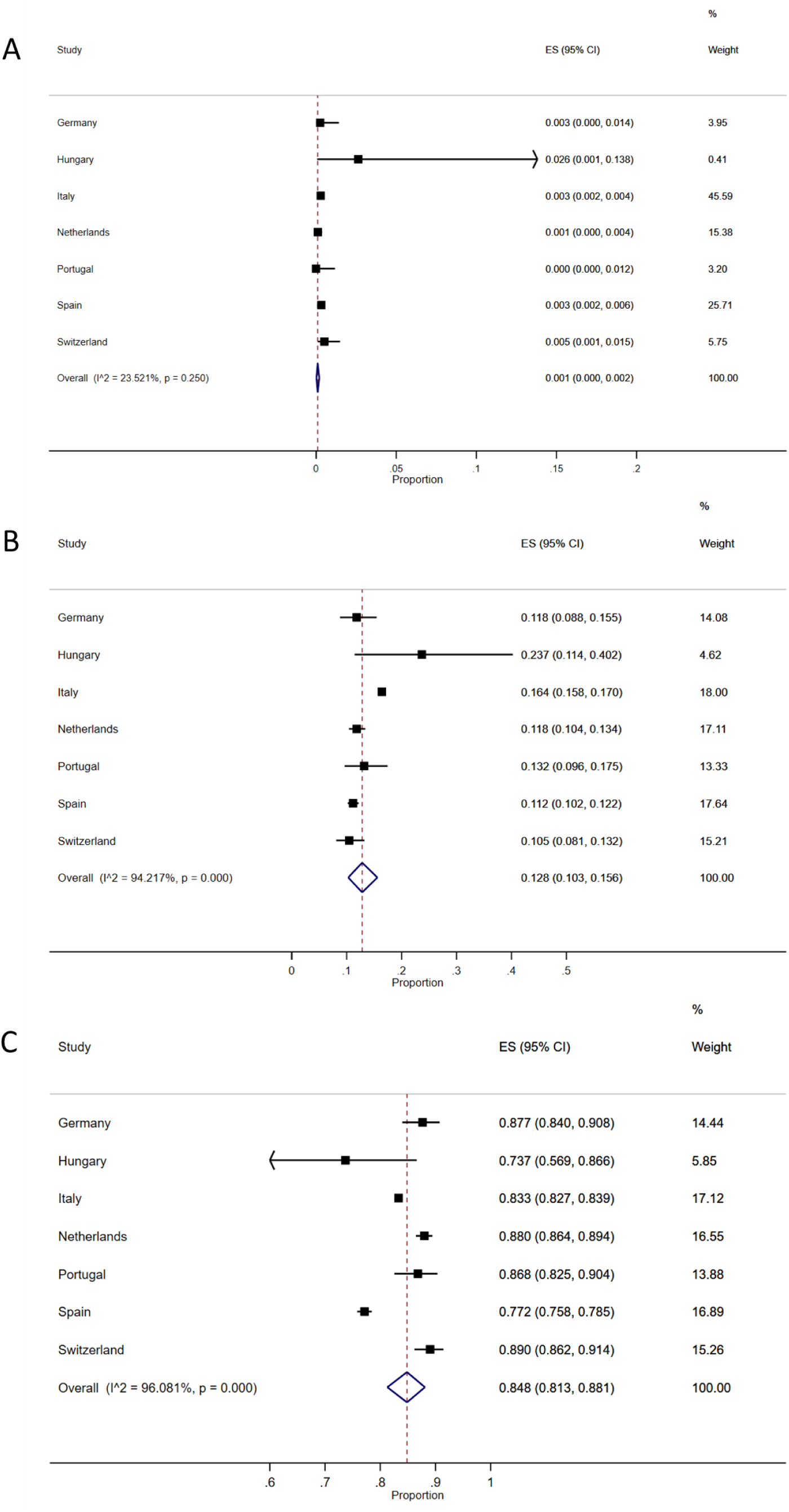
Distribution of age groups among all COVID-19-related deaths in Europe: meta-analysis. (A) : Patients < 40 years; (B): 40-69 years; (C): ≥ 70 years of age.

## Discussion

This report describes the current distribution of COVID-19-related deaths in Europe across age groups, using official mortality data from 13 countries. This represents about half of the total of COVID-19-related deaths reported worldwide as of April 6, 2020 [1]. We found that persons under 40 years represent about 0.1% of COVID-19-related deaths, while persons over 70 years represent about 85% of fatalities.

The distribution of COVID-19-related deaths by age in Europe differs from what was observed in the early phase of the pandemic in mainland China (as of February 11, 2020: <40, 40-69, ≥ 70 years old representing 2.5%, 46.6%, and 50.8% of 1,023 COVID-19-related deaths, respectively) [3], and of what is currently observed in the US (as of April 7: <45, 45-64, ≥ 65 years old representing 3.3%, 17.6%, and 79.0% of 2,214 COVID-19-related deaths, respectively) [9]. This could reflect different patterns in patient characteristics, underlying risk factors, and patient management across settings, as well as variability in the organization of healthcare and identification of causes of death across countries. The age structure of each population might also be key: in 2019, Italy had the highest proportion of population aged ≥ 80 years in Europe (7.2%), while in the US, for example, this proportion was as low as 3.6% in 2010 [10]. On the other hand, the overall prevalence of obesity (body mass index ≥ 30), which has been reported as a risk factor for severe COVID-19, is much higher in the US compared to Italy (42.4% vs. 10.5%, respectively) [11].

Our study has limitations. First, we could not include all European countries, mainly because official reports on COVID-19 related mortality data by age group were not always available. Second, we could not investigate the potential burden of underlying health conditions and other risk factors such as obesity and chronic lung disease because this information was rarely notified. Third, we investigated COVID-19-related mortality data, but we acknowledge that methods for case ascertainment, case definitions, and SARS-CoV-2 RT-PCR testing strategies varied across countries. Some countries, such as France, provided detailed information only for in-hospital fatalities, which does not account for COVID-related deaths that occurred in other settings such as nursing homes, for a potential overrepresentation of younger patients. Fourth, we focused on COVID-19-related deaths, but future studies should also examine the distribution of age among patients with COVID-19 admitted to intensive care units.

Physical distancing is currently recommended in many countries for all age groups to slow the spread of COVID-19 and protect the elderly and, more broadly, the healthcare system. People under 40 years of age represent a very small fraction of the total number of COVID-19-related deaths in Europe. These results, together with evaluations of the impact of comorbidities and risk factors in the course of COVID-19, may help health authorities respond to public concerns and guide future physical distancing and mitigation strategies.

## Data Availability

The data is publicly available.

## Supplementary Materials

None.

## Author Contributions

Conceptualization, J.F.C., M.C., J.T.; Methodology, J.F.C. and M.T.; Statistical analysis, J.F.C.; Data extraction and management, J.F.C. and D.A.K.; Writing—original draft preparation, J.F.C. and J.T.; Writing—review and editing, J.F.C., D.A.K., S.M., J.B., M.C., J.T.; Supervision, MC and J.T. All authors have read and agreed to the published version of the manuscript.

## Acknowledgments

We thank Prof. Richard Malley (Boston Children’s Hospital, USA) and Prof. Henrique Barros (Institute of Public Health, University of Porto, Portugal) for their valuable feedback on the manuscript.

## Appendices

## Appendix 1. Sources of data (all websites accessed April 6, 2020)

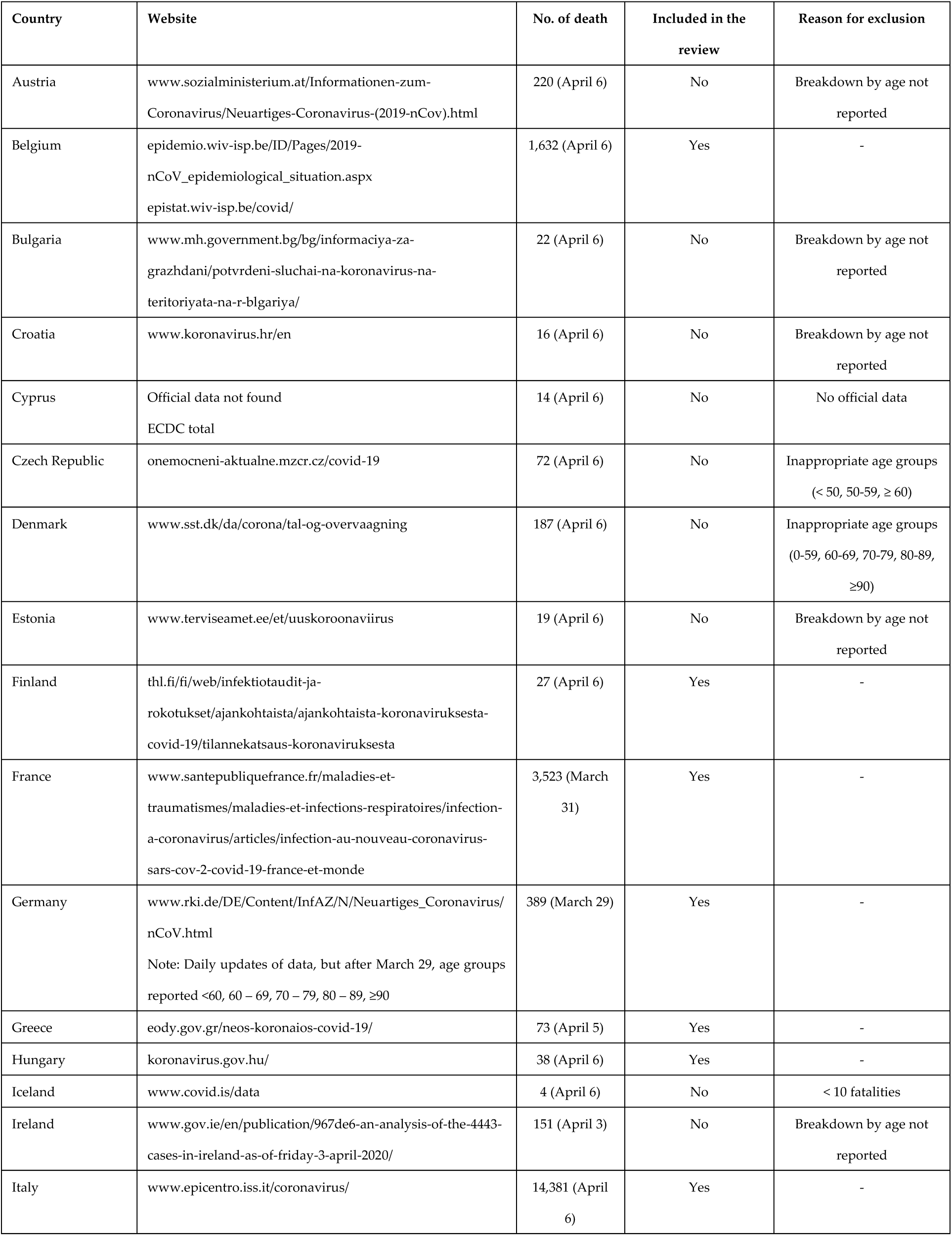
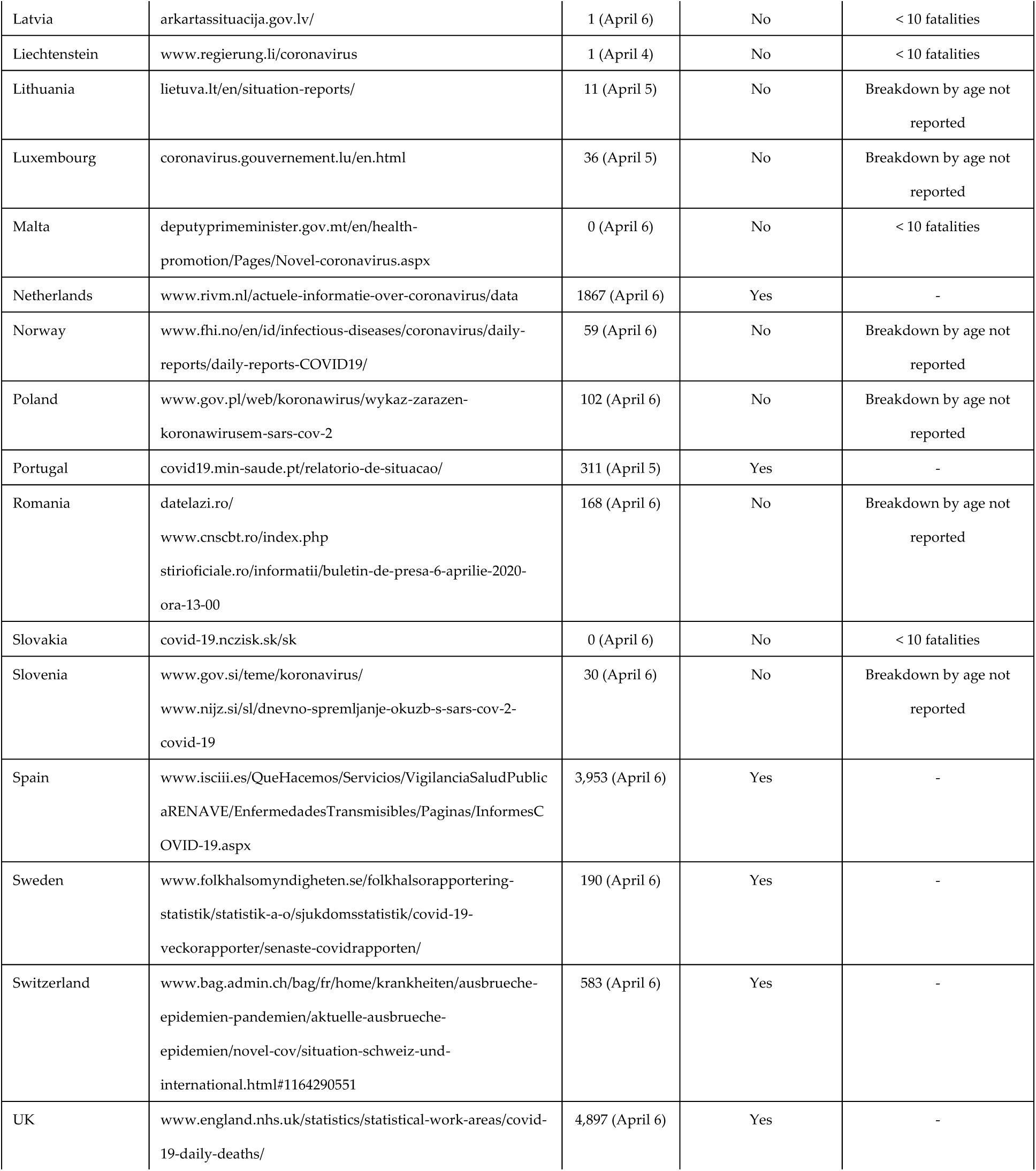

## Appendix 2. Sensitivity analysis including countries with less stringent age groups Tables and figures

Appendix 2. Sensitivity analysis including countries with less stringent age groups A. Youngest age group: around <40-50
| Country | Prop | [95% Conf. Interval] |  | % Weight |
| --- | --- | --- | --- | --- |
| Belgium | 0.006 | 0.003 | 0.010 | 10.84 |
| Finland | 0.000 | 0.000 | 0.128 | 0.61 |
| France | 0.008 | 0.006 | 0.012 | 12.82 |
| Germany | 0.003 | 0.000 | 0.014 | 5.65 |
| Greece | 0.014 | 0.000 | 0.074 | 1.53 |
| Hungary | 0.026 | 0.001 | 0.138 | 0.84 |
| Italy | 0.003 | 0.002 | 0.004 | 14.56 |
| Netherlands | 0.001 | 0.000 | 0.004 | 11.25 |
| Portugal | 0.000 | 0.000 | 0.012 | 4.88 |
| Spain | 0.003 | 0.002 | 0.006 | 13.05 |
| Sweden | 0.005 | 0.000 | 0.029 | 3.41 |
| Switzerland | 0.005 | 0.001 | 0.015 | 7.14 |
| UK | 0.009 | 0.006 | 0.012 | 13.42 |
| Random pooled ES | 0.002 | 0.001 | 0.004 | 100.00 |

B. Intermediate age group: around 40-69 years
| Country | Prop | [95% Conf. Interval] |  | % Weight |
| --- | --- | --- | --- | --- |
| Belgium | 0.069 | 0.057 | 0.082 | 9.02 |
| Finland | 0.074 | 0.009 | 0.243 | 3.30 |
| France | 0.091 | 0.081 | 0.101 | 9.17 |
| Germany | 0.118 | 0.088 | 0.155 | 8.24 |
| Greece | 0.247 | 0.153 | 0.361 | 5.54 |
| Hungary | 0.237 | 0.114 | 0.402 | 4.05 |
| Italy | 0.164 | 0.158 | 0.170 | 9.27 |
| Netherlands | 0.118 | 0.104 | 0.134 | 9.06 |
| Portugal | 0.132 | 0.096 | 0.175 | 8.02 |
| Spain | 0.112 | 0.102 | 0.122 | 9.18 |
| Sweden | 0.263 | 0.202 | 0.332 | 7.37 |
| Switzerland | 0.105 | 0.081 | 0.132 | 8.57 |
| UK | 0.072 | 0.065 | 0.080 | 9.21 |
| Random pooled ES | 0.125 | 0.098 | 0.155 | 100.00 |

C. Oldest age group: around ≥60-70 years
| Country | Prop | [95% Conf. Interval] |  | % Weight |
| --- | --- | --- | --- | --- |
| Belgium | 0.912 | 0.898 | 0.926 | 8.89 |
| Finland | 0.926 | 0.757 | 0.991 | 3.62 |
| France | 0.888 | 0.877 | 0.898 | 9.01 |
| Germany | 0.877 | 0.840 | 0.908 | 8.23 |
| Greece | 0.740 | 0.624 | 0.835 | 5.81 |
| Hungary | 0.737 | 0.569 | 0.866 | 4.37 |
| Italy | 0.833 | 0.827 | 0.839 | 9.09 |
| Netherlands | 0.880 | 0.864 | 0.894 | 8.92 |
| Portugal | 0.868 | 0.825 | 0.904 | 8.04 |
| Spain | 0.772 | 0.758 | 0.785 | 9.02 |
| Sweden | 0.732 | 0.663 | 0.793 | 7.47 |
| Switzerland | 0.890 | 0.862 | 0.914 | 8.51 |
| UK | 0.919 | 0.911 | 0.927 | 9.04 |
| Random pooled ES | 0.855 | 0.821 | 0.886 | 100.00 |

$I^2$ (variation in ES attributable to heterogeneity) = 97.9%

